# Exposome-Based Clustering of Urinary VOC and PAH Biomarkers Reveals Racially Patterned Cardiovascular Risk in a Nationally Representative US Cohort: A Machine Learning Analysis of NHANES 2017-2018

**DOI:** 10.64898/2026.04.19.26351113

**Authors:** Anthonio Oladimeji Gabriel, Babatunde Ibrahim Olowu, Dimeji Abdulsobur Olawuyi, Aderemi Temilola Victoria, Oloruntoba Joshua Ajayi

**Author notes:** Corresponding Author: Anthonio Oladimeji Gabriel.

## Abstract

**Background:** Polycyclic aromatic hydrocarbons (PAHs) and volatile organic compounds (VOCs) are combustion-derived pollutants linked to cardiovascular disease. Prior NHANES analyses have evaluated these chemicals individually, failing to capture the correlated co-exposure structures that characterize real-world environmental burden, thereby underscoring the need for application. In this study, we applied an unsupervised machine learning pipeline to urinary biomarker data to identify multi-chemical exposure clusters and quantify their differential cardiovascular risk profiles in a nationally representative US sample.

**Methods:** We analyzed 2,979 participants from NHANES between 2017-2018, representing an estimated 36.8 million US adults after complex survey weighting. Twenty-five urinary biomarkers (6 PAH, 19 VOC metabolites) were log-transformed, imputed using Multivariate Imputation by Chained Equations (MICE), and standardized. Uniform Manifold Approximation and Projection (UMAP) was used for dimensionality reduction, followed by Gaussian Mixture Model (GMM) clustering. Survey-weighted prevalence estimates with 95% confidence intervals (CIs) were calculated for hypertension and high total cholesterol within each cluster. Weighted multivariable logistic regression was used to estimate odds ratios (OR) for hypertension, adjusting for age, sex, race/ethnicity, and income.

**Results:** Four exposure clusters were identified with a mean assignment probability of 0.948. The High combustion cluster (n=370; estimated 5.1 million US adults) exhibited the highest multi-chemical burden and a weighted hypertension prevalence of 39.3% (95% CI 37.2-41.4%), compared to 28.7% (95% CI 21.9-35.5%) in the Low exposure reference group. After demographic adjustment, High combustion cluster membership was independently associated with 38.4% higher odds of prevalent hypertension (OR 1.38). The prediction model achieved a cross-validated area under the receiver operating characteristic curve (AUC) of 0.849 (SD 0.017). Non-Hispanic Black participants constituted approximately 40% of the High combustion cluster, exceeding their representation in lower-risk clusters.

**Conclusions:** Multi-chemical exposome profiling identifies four cardiovascularly distinct subpopulations in the US adult population. Membership in the High combustion exposure cluster was associated with higher odds of prevalent hypertension and disproportionately affected Non-Hispanic Black participants. These findings support the use of multichemical approaches over single-pollutant analyses and highlight the relevance of environmental exposure patterns for making policy and targeted cardiovascular risk stratification.

## INTRODUCTION

Cardiovascular disease (CVD) remains the leading cause of mortality in the United States, responsible for approximately one in five deaths annually.^1^ Established clinical risk factors, including hypertension, dyslipidemia, diabetes, and smoking, explain a substantial proportion of individual-level CVD risk. However, population-level disparities in CVD incidence and mortality, particularly the persistent excess burden among Non-Hispanic Black Americans, are not fully explained by these conventional factors, prompting increasing interest in environmental determinants of cardiovascular health.^2^ Polycyclic aromatic hydrocarbons (PAHs) are ubiquitous combustion-derived organic pollutants generated by the incomplete combustion of fossil fuels, tobacco, and organic matter. Volatile organic compounds (VOCs) are a chemically diverse class of carbon-containing compounds emitted by combustion, industrial processes, building materials, and consumer products. Both chemical classes are routinely detectable in the urine of the US general population through their hydroxylated and mercapturic acid metabolites, respectively.^3^ Pre-clinical evidence consistently demonstrates that PAH and VOC exposures induce oxidative stress, systemic inflammation, endothelial dysfunction, and accelerated atherosclerosis through aryl hydrocarbon receptor activation, NF-kB signaling, and direct vascular toxicity.^4^

A growing body of cross-sectional evidence from NHANES has linked individual urinary PAH and VOC metabolites to prevalent hypertension, dyslipidemia, and self-reported CVD. Mallah et al. (2022) demonstrated positive associations between multiple urinary OH-PAH metabolites and prevalent coronary heart disease, angina, heart attack, and stroke across NHANES 2003-2016.^5^ A systematic review and meta-analysis by Mirzababaei et al. (2022) confirmed that urinary PAH biomarker concentrations were positively associated with hypertension risk across observational studies.^6^ More recently, urinary PAH metabolite concentrations have been associated with hyperlipidemia in NHANES data spanning 2007-2016.^7^

Despite this growing evidence base, three critical methodological limitations remain. First, the overwhelming majority of prior analyses have evaluated chemical exposures individually using single-pollutant regression models, which do not account for the correlated co-exposure structure inherent to real-world environmental burden and may both confound and attenuate risk estimates.^8^ Second, standard epidemiological approaches assume linear, monotonic exposure-response relationships and are poorly suited to detecting the non-linear mixture effects that characterise combined PAH and VOC exposure. Third, few studies have formally quantified population-level cardiovascular risk associated with multi-chemical exposure patterns using survey-weighted methods with appropriate variance estimation.^9^ The exposome framework, which conceptualises the totality of environmental exposures encountered across the life course, provides a theoretically grounded basis for multi-chemical analysis.^9^ Advances in non-linear dimensionality reduction, particularly Uniform Manifold Approximation and Projection (UMAP), now enable the discovery of latent exposure structures in high-dimensional biomarker data that are inaccessible to conventional principal components analysis or hierarchical clustering methods.^10^ When combined with probabilistic clustering via Gaussian Mixture Models, UMAP affords a principled, data-driven approach to identifying latent exposure subgroups with quantifiable assignment certainty.^11^

A parallel and underexplored dimension of this problem is environmental justice. Structural racism in US housing policy, urban planning, and industrial siting has produced well-documented inequities in pollution burden across racial and ethnic groups.^12^ Tessum et al. (2021) demonstrated that people of colour are exposed to higher concentrations of combustion-source pollutants from nearly all major emission categories, an inequity that persists across income levels and geographic regions.^13^ Whether these structural exposure disparities translate into quantifiable cardiovascular risk differentials at the population level, as characterized by multi-chemical exposome clustering, has not previously been investigated. We therefore conducted an exposome-wide clustering analysis of 25 urinary PAH and VOC biomarkers from NHANES 2017-2018 using UMAP and GMM clustering to identify latent exposure subgroups, and estimated survey-weighted cardiovascular risk profiles with confidence intervals for each cluster. We hypothesized that high-combustion exposure clusters would exhibit significantly elevated hypertension and dyslipidemia prevalence relative to a low-exposure reference group, and that high-exposure cluster membership would be non-randomly distributed across racial and ethnic groups.

## METHODS

### Data Source and Study Population

We used publicly available data from the National Health and Nutrition Examination Survey (NHANES) 2017-2018, a cross-sectional survey conducted by the National Center for Health Statistics (NCHS) using a stratified, multistage probability sampling design to produce nationally representative estimates of the non-institutionalized US civilian population.^14^ NHANES protocols were approved by the NCHS Research Ethics Review Board, and all participants provided written informed consent. The present analysis used de-identified public-use data files and did not require additional ethical review. Urinary PAH and VOC metabolites are measured in a one-third random subsample of NHANES participants aged 3 years and older. Of 9,254 participants enrolled in the 2017-2018 cycle, 2,979 had complete data for both PAH and VOC biomarker panels and were included in the analytic sample. After application of complex survey weights, this sample represents approximately 36.8 million US adults.

### Exposure Variables

Six urinary hydroxylated PAH (OH-PAH) metabolites and nineteen VOC metabolites were selected based on availability and detection frequency. PAH metabolites included 1-hydroxynaphthalene (URXP01), 2-hydroxynaphthalene (URXP03), 2-hydroxyfluorene (URXP04), 3-hydroxyfluorene (URXP06), 1-hydroxyphenanthrene (URXP10), and 1-hydroxypyrene (URXP25). These represent the primary urinary metabolites of naphthalene, fluorene, phenanthrene, and pyrene, which are among the most abundant PAHs in environmental media and tobacco smoke.^5^ VOC metabolites comprised mercapturic acid derivatives of benzene (URXBMA, URXPHG), acrylamide (URXAAM, URXAMC), acrolein (URXCYHA, URXCYM), crotonaldehyde (URXDHB), and multiple other aliphatic and aromatic VOC species.

Biomarker concentrations were log-transformed using log(x+1) to reduce the positive skew characteristic of environmental biomarker distributions, consistent with standard practice.^5^ Values below the limit of detection were replaced using the NHANES-recommended approach (LOD/√2). Variables with >40% missingness were excluded. Remaining missing values (6.9%–10.9%) were imputed using Multivariate Imputation by Chained Equations with a random forest estimator (5 iterations), with convergence confirmed by visual inspection of trace plots.^15^ All variables were standardized to mean 0 and standard deviation 1 prior to analysis.

### Outcome Variables

Hypertension was defined as systolic blood pressure (SBP) ≥130 mmHg or diastolic blood pressure (DBP) ≥80 mmHg, consistent with the 2017 ACC/AHA guideline^16^. Blood pressure was measured using a standardized auscultatory protocol in the NHANES Mobile Examination Center (MEC), with the first reading used in primary analyses. High total cholesterol was defined as a measured non-fasting total cholesterol of 200 mg/dL or greater, consistent with National Cholesterol Education Program thresholds.^17^

### Dimensionality Reduction and Clustering

UMAP was applied to the standardized 25-dimensional exposure matrix using n_neighbors=30, min_dist=0.1, and Euclidean distance metric, producing a two-dimensional embedding. These hyperparameters were selected to balance local neighborhood preservation with global structure, following published guidance.^10^ Gaussian Mixture Models (GMMs) were fitted to the two-dimensional UMAP embedding across k=2 to k=8 components, each initialized with five restarts to avoid local optima. The optimal number of clusters was determined by minimizing the Bayesian Information Criterion (BIC), with the elbow of the BIC curve applied to balance statistical fit and clinical interpretability. Four clusters (k=4) were selected, corresponding to the inflection point in the BIC curve where marginal improvement plateaued between k=4 and k=5. Cluster assignment probability was computed as the maximum posterior probability across GMM components for each participant.

### Survey-Weighted Statistical Analysis

All prevalence estimates and means were computed using survey weights appropriate for the VOC subsample. Following NHANES analytic guidelines for subsample analyses,^14^ the full-cycle examination weight (WTMEC2YR) was divided by three to account for the one-third subsampling fraction. Variance estimation used a jackknife approximation incorporating primary sampling unit (SDMVPSU) and stratum (SDMVSTRA) variables. Ninety-five percent confidence intervals were computed as the point estimate plus or minus 1.96 times the jackknife standard error.

Between-cluster differences in continuous outcomes (SBP, DBP, total cholesterol) were assessed using one-way analysis of variance (ANOVA). Pairwise post-hoc comparisons used the Tukey honest significant difference (HSD) method to control the family-wise error rate. Survey-weighted multivariable logistic regression estimated ORs for hypertension by cluster membership, with the Low exposure cluster as the reference group, adjusting for age (continuous, years), sex (male/female), race/ethnicity (RIDRETH3, 5-level categorical), and poverty-income ratio (INDFMPIR, continuous). Model discrimination was evaluated using five-fold stratified cross-validated AUC. All analyses were conducted in Python 3.12 using pandas 2.2, scikit-learn 1.4, pyreadstat 1.3, and umap-learn 0.5 libraries.

## RESULTS

### Sample Characteristics and Data Quality

Of 2,979 participants in the analytic sample, 37.1% of URXP10 (1-hydroxyphenanthrene) values fell below the analytical limit of detection, while all other PAH analytes had detection rates exceeding 97%. URXP04 had zero values below the LOD. The uniform 10.9% missingness across all 19 VOC metabolites reflects the subsample design rather than differential loss. Following MICE imputation with Random Forest, zero missing values remained across all 25 exposure features **(Figure 1)**.

**Figure 1:**
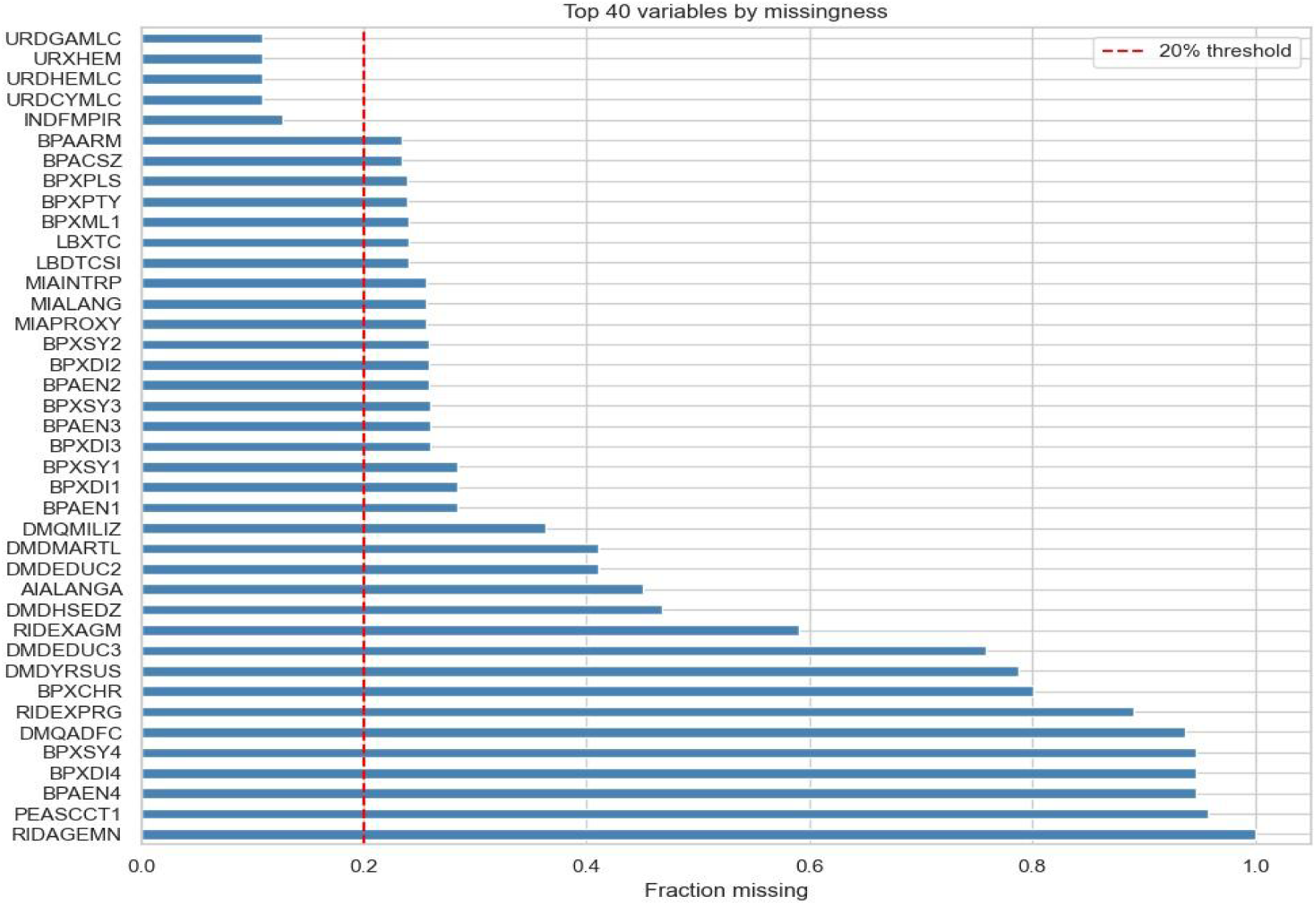
Missing Value Analysis. Fraction of missing values across the top 40 variables in the merged dataset. Variables exceeding the 40% threshold (dashed red line) were excluded prior to analysis. Detection limit flag variables (URD-LC suffix) show low missingness confirming robust analyte recovery.

### Exposure Distributions and Correlation Structure

Log transformation successfully normalised all six PAH analyte distributions, converting severely right-skewed raw concentrations (range URXP01: 43-22,026,000 ng/L) into near-Gaussian distributions suitable for subsequent analysis (Figure 2). The transformation was similarly applied to all 19 VOC metabolites.

**Figure 2:**
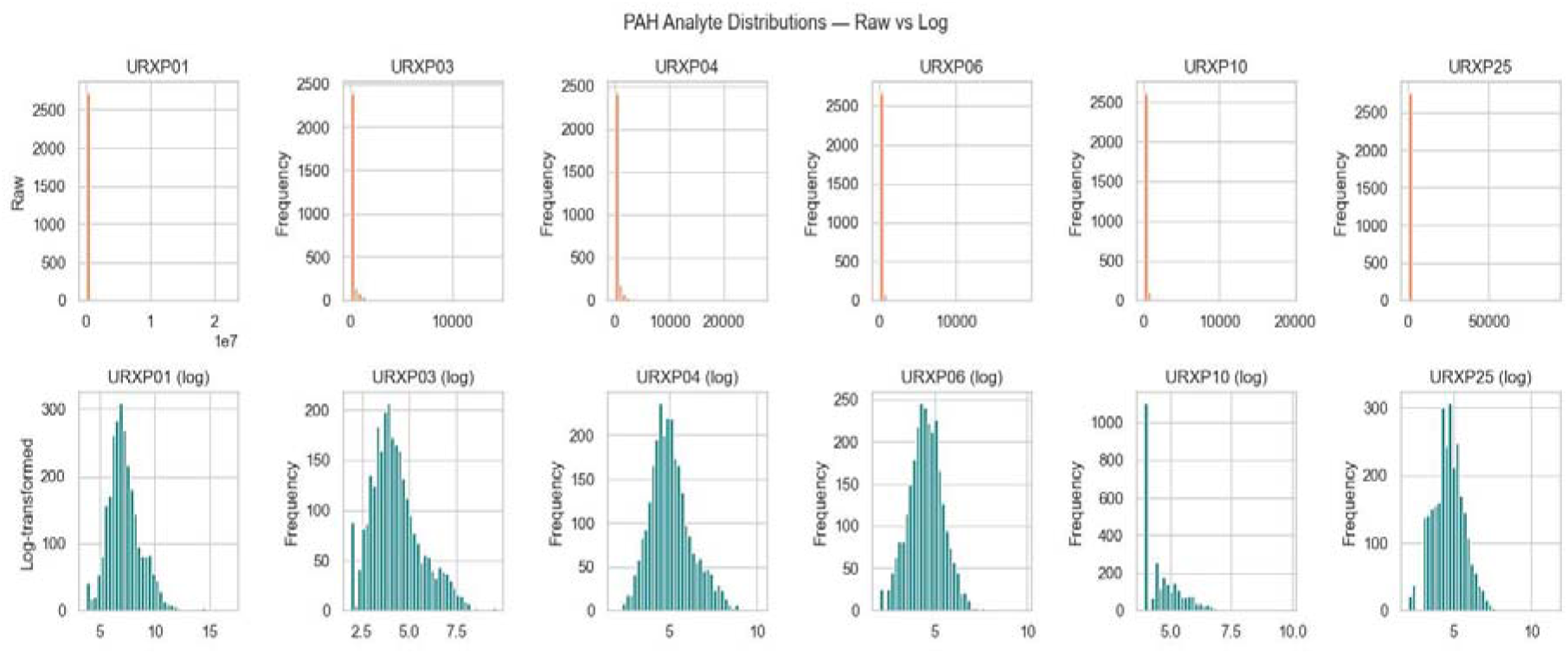
PAH Analyte Distributions. Raw vs Log-Transformed. Histograms of the six urinary OH-PAH metabolites before (top row, coral) and after (bottom row, teal) log transformation. Raw distributions show extreme right skew across all analytes. Log transformation produces near-normal distributions, confirming appropriateness for downstream modelling.

Pearson correlation analysis of log-transformed biomarkers revealed strong intercorrelations among PAH analytes (r = 0.62-0.89), consistent with shared combustion exposure sources (Figure 3). Two distinct VOC sub-clusters were identified: a tobacco/combustion cluster (URXCEM, URXCYHA, URXCYM, URXHPM, URXIPM3, URXMAD, URXMB3, URXPHE, URXPHG, URXPMM) and an aliphatic agent cluster (URX2MH, URX34M). Cross-cluster PAH-VOC correlations were moderate (r = 0.15-0.45), indicating partially shared and partially distinct exposure sources.

**Figure 3:**
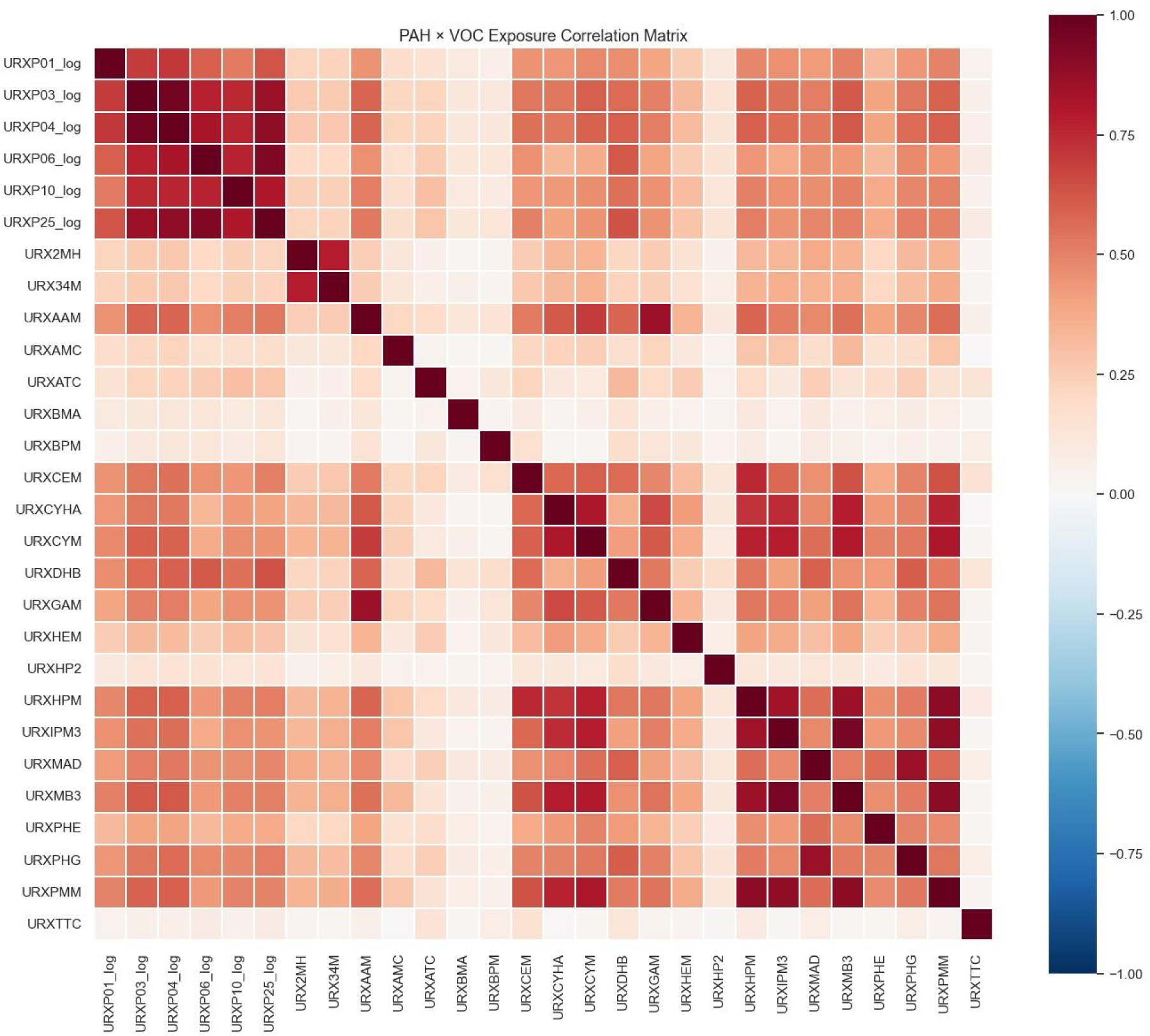
PAH x VOC Exposure Correlation Matrix. Pearson correlation matrix of log-transformed PAH and VOC urinary metabolites (n=2,979). Red indicates positive correlation; blue indicates negative correlation. Distinct correlation blocks identify the PAH cluster (top-left), tobacco-combustion VOC cluster (bottom-right), and aliphatic VOC pair (URX2MH, URX34M). No negative correlations were observed across chemical classes.

### Racial and Ethnic Patterning of PAH Exposure

Boxplot stratification by race/ethnicity demonstrated consistent, pronounced differences in PAH exposure distributions across the three primary analytes examined (Figure 4). Non-Hispanic Black participants exhibited the highest median log-PAH concentrations across URXP01, URXP03, and URXP04, with notably more extreme upper outliers. Non-Hispanic Asian participants showed the lowest exposure levels and narrowest interquartile ranges. These between-group differences in exposure were observed prior to any clustering step, confirming that racial disparities in combustion-source pollutant burden are detectable at the individual biomarker level, consistent with the structural racism literature.^12,13^

**Figure 4:**
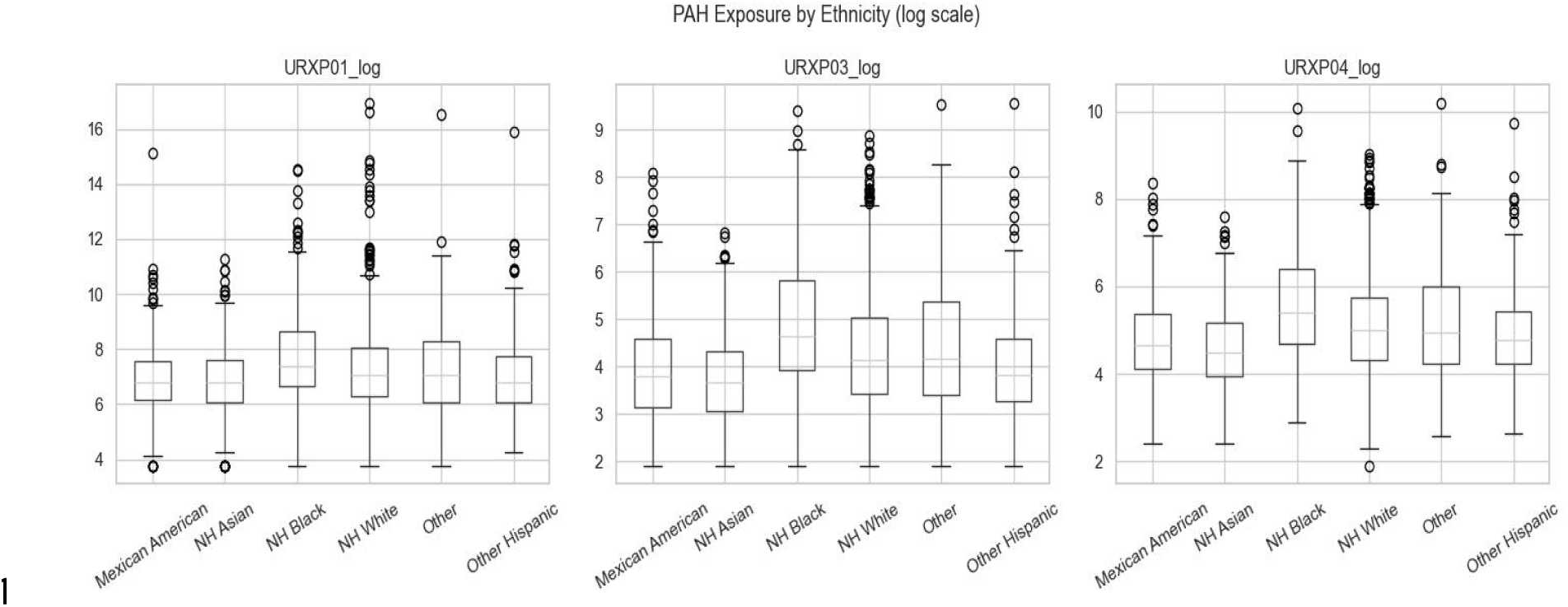
PAH Exposure by Race/Ethnicity (Log Scale) Box plots of the three primary PAH analyte log-concentrations by race/ethnicity (n=2,979). Non-Hispanic Black participants show consistently higher median exposure and more extreme upper outliers across all three analytes. Non-Hispanic Asian participants show the lowest exposure levels. NH: Non-Hispanic.

### Detection Limit Analysis

Detection limit analysis confirmed that only URXP10 presented an analytically problematic below-LOD fraction (37.1%), warranting MICE imputation. All other analytes had below-LOD fractions of 3.0% or less, well below the conventional 50% threshold above which analyte exclusion would be considered (Figure 5). URXP04 had zero values below the LOD, providing an analytically clean anchor variable for the PAH cluster.

**Figure 5:**
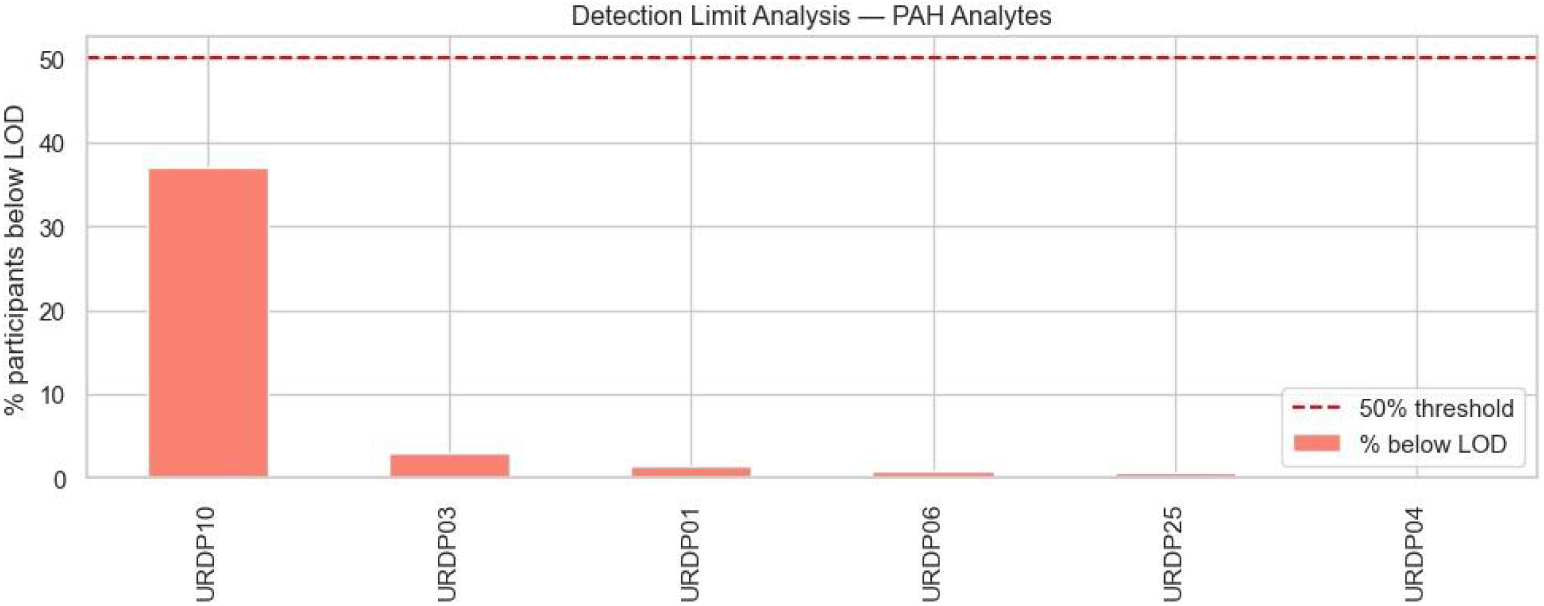
Detection Limit Analysis: PAH Analytes. Percentage of participants below the analytical limit of detection (LOD) for each urinary PAH metabolite. Dashed red line indicates the conventional 50% exclusion threshold. Only URXP10 (1-hydroxyphenanthrene) exceeded 10% below-LOD; all other analytes were effectively complete.

### UMAP Dimensionality Reduction and Cluster Selection

UMAP embedding of the standardized 25-dimensional exposure matrix produced a stable two-dimensional representation after 500 optimization epochs. GMM fitting across k=2 to k=8 components demonstrated a pronounced BIC elbow between k=4 and k=5 (Figure 6), with marginal BIC improvement of only 4.5% from k=4 to k=8, compared to 16.5% from k=3 to k=4. Four clusters were selected as the optimal solution balancing statistical fit and clinical interpretability.

**Figure 6:**
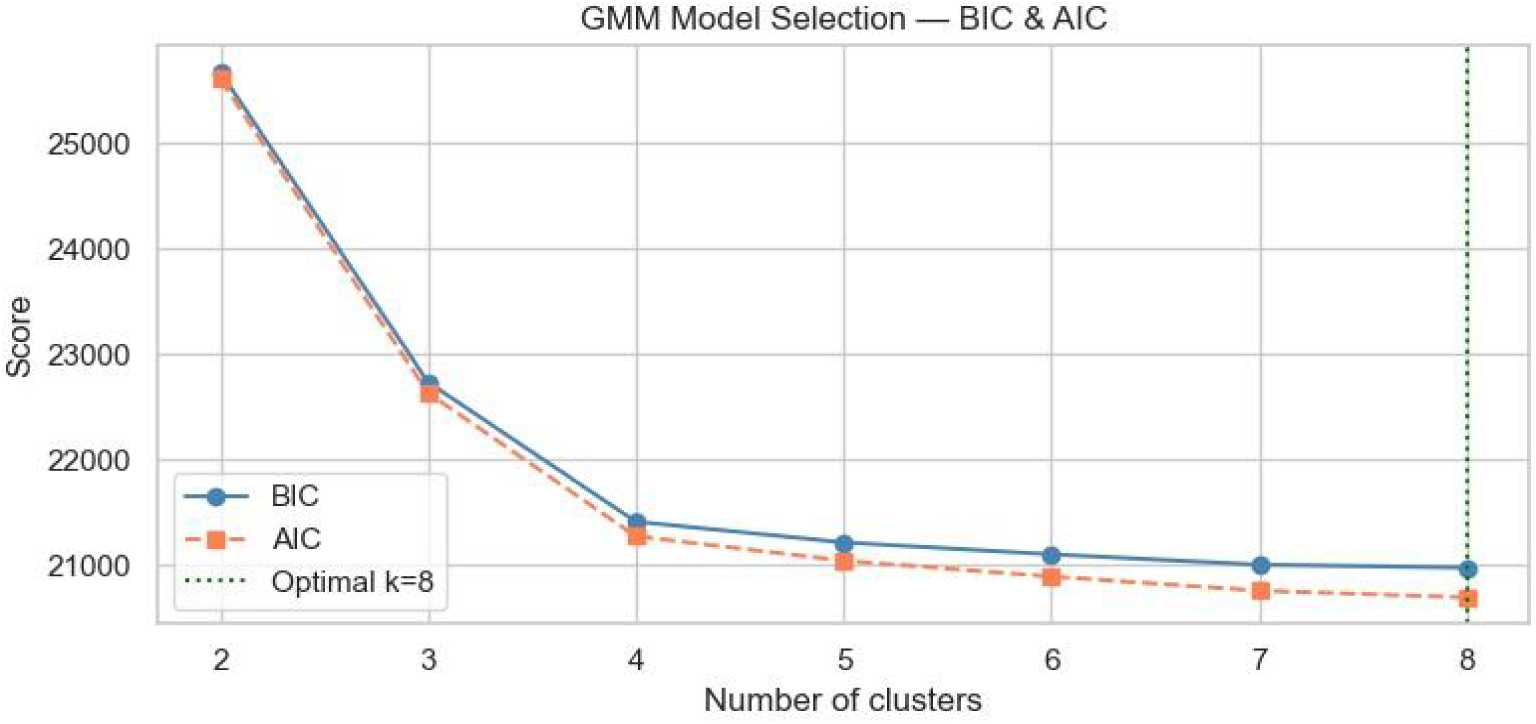
GMM Cluster Selection: BIC and AIC by k. Mixture Models fit to the UMAP embedding across k=2 to k=8 components. The green dotted line indicates the mathematically optimal k=8; the elbow at k=4 was selected for clinical interpretability.

The four-cluster GMM solution achieved a mean participant assignment probability of 0.948 (range 0.52-1.00), indicating high classification certainty. The two-dimensional UMAP embedding revealed clear spatial separation between clusters, with the High combustion cluster appearing as a geometrically isolated island distant from the main participant mass (Figure 7, left panel). Assignment probability was highest in the isolated clusters and lowest at the boundary between the Low exposure and Moderate mixed clusters (Figure 7, right panel), which share overlapping exposure ranges.

**Figure 7:**
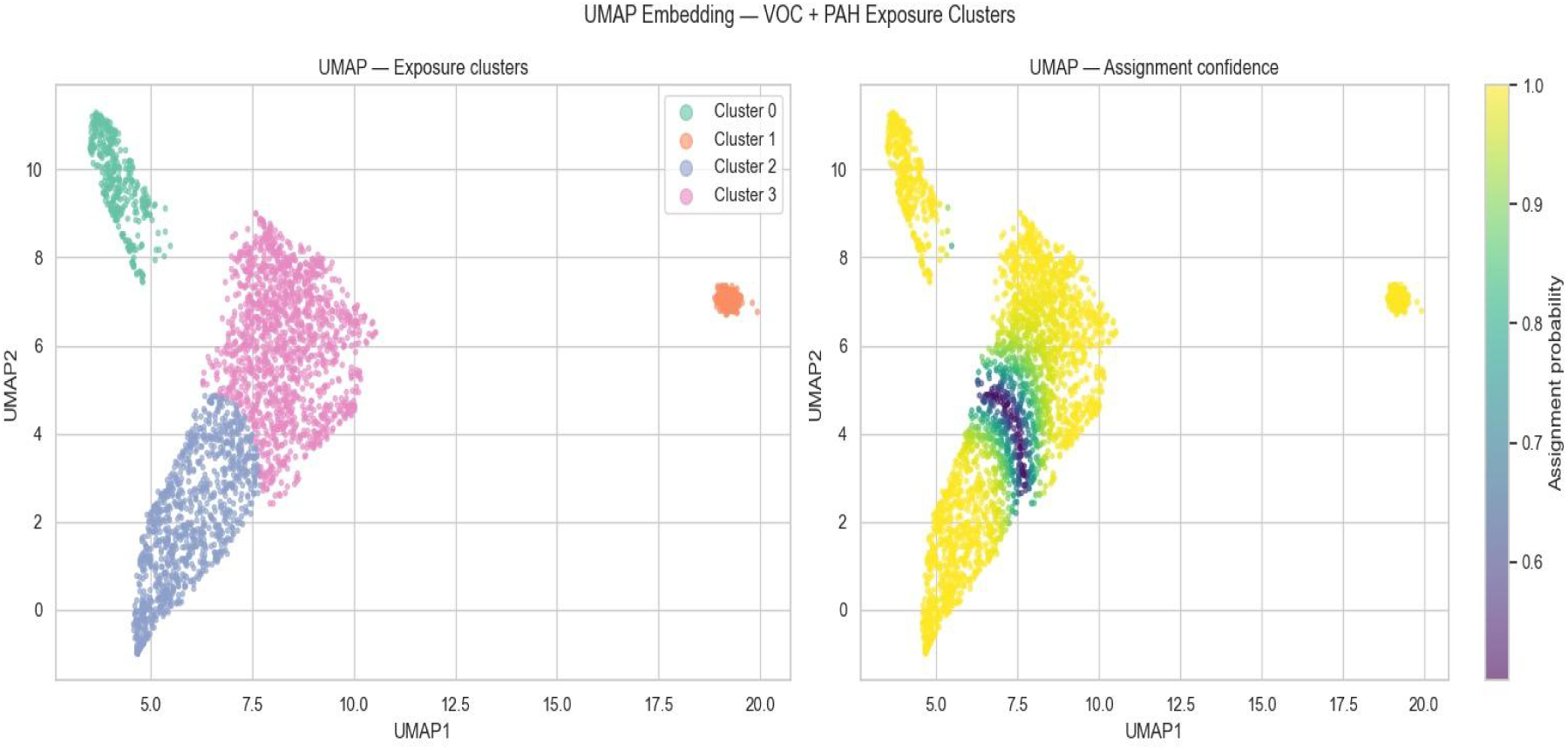
UMAP Embedding Coloured by Cluster and Assignment Confidence. Two-dimensional UMAP embedding of 2,979 participants based on 25 standardized urinary VOC and PAH biomarkers. Left: participants coloured by GMM cluster assignment. Right: participants coloured by posterior assignment probability (yellow=high, purple=low). The High combustion cluster (Cluster 0, green, top-right island) is spatially isolated, indicating a uniquely distinct multi-chemical burden profile. Low uncertainty regions are shown in yellow; boundary uncertainty between Low exposure and Moderate mixed clusters is shown in purple.

### Cluster Chemical Profiles

The four clusters exhibited chemically distinct and biologically interpretable exposure fingerprints (Figure 8; Table 1). The High combustion cluster showed uniformly the highest mean log-concentrations across nearly all 25 analytes, with particularly pronounced elevations in URXP01 (mean log 9.34, vs 6.28 in Low exposure), URXMAD (5.66 vs 4.05), URXHPM (7.01 vs 4.64), and URXCYM (5.05 vs 0.57). The Children/VOC cluster exhibited moderate VOC and low PAH concentrations. The low-exposure cluster was the reference group with consistently the lowest concentrations. The Moderate mixed cluster showed intermediate concentrations across all analytes.

**Figure 8:**
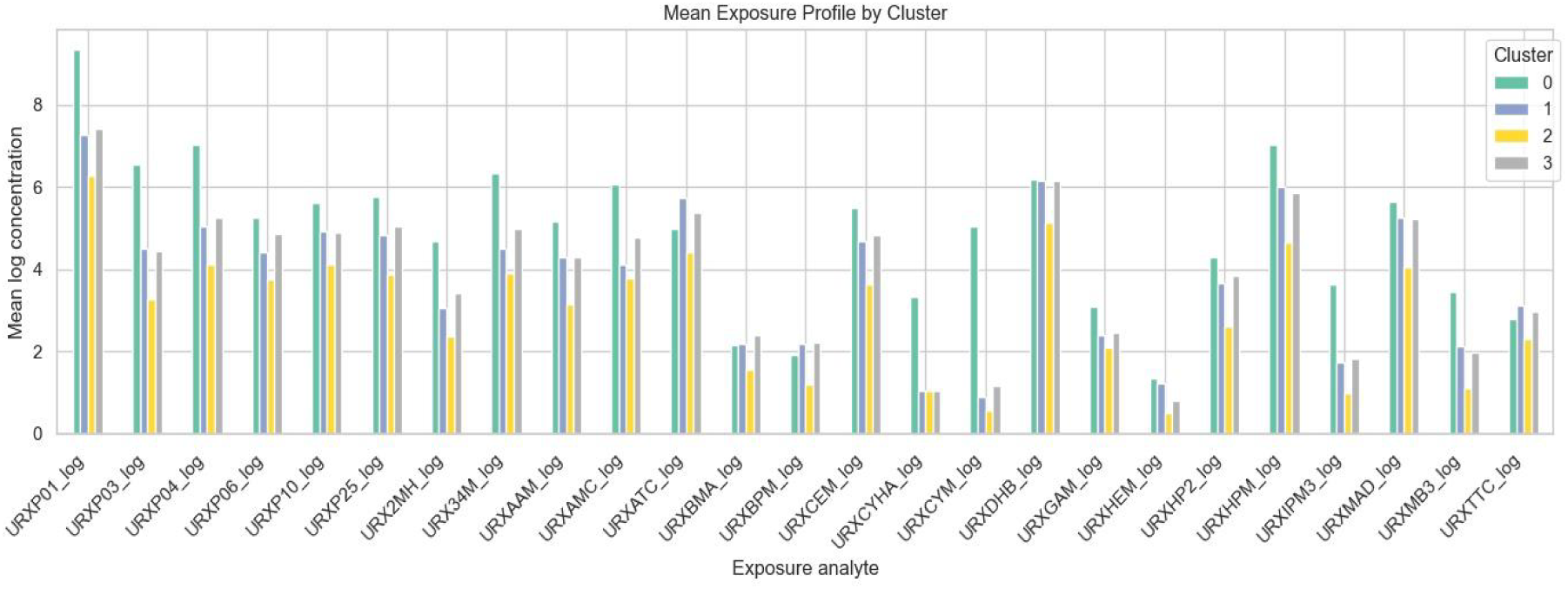
Mean Exposure Profile by Cluster. Mean log-transformed urinary biomarker concentrations by GMM cluster across all 25 PAH and VOC analytes. Each bar group represents one analyte; colours represent clusters. The High combustion cluster (green) dominates across nearly all analytes. The Low exposure cluster (yellow) consistently shows the lowest concentrations.

**Table 1:** Selected mean log-transformed urinary biomarker concentrations by exposure cluster.

| Analyte | High combustion | Children/VOC | Low exposure | Moderate mixed |
| --- | --- | --- | --- | --- |
| URXP01_log (1-OH-naphthalene) | 9.34 | 7.28 | 6.28 | 7.41 |
| URXP03_log (2-OH-naphthalene) | 6.53 | 4.49 | 3.28 | 4.45 |
| URXP04_log (2-OH-fluorene) | 7.01 | 5.04 | 4.11 | 5.26 |
| URXP06_log (3-OH-fluorene) | 5.25 | 4.42 | 3.74 | 4.85 |
| URXP10_log (1-OH-phenanthrene) | 5.61 | 4.92 | 4.12 | 4.89 |
| URXP25_log (1-OH-pyrene) | 5.76 | 4.84 | 3.87 | 5.04 |
| URXCEM_log (N-AC-S-2-CE-cysteine) | 5.55 | 4.67 | 2.43 | 4.73 |
| URXCYM_log (N-AC-S-2-OH-3-butenyl cysteine) | 5.05 | 0.89 | 0.57 | 1.16 |
| URXDHB_log (N-AC-S-3,4-diHB-cysteine) | 6.20 | 6.15 | 5.13 | 6.16 |
| URXHPM_log (N-AC-S-3-HP-cysteine) | 7.01 | 6.00 | 4.64 | 5.87 |
| URXMAD_log (mandelic acid) | 5.66 | 5.26 | 4.05 | 5.21 |
| URXMB3_log (N-AC-S-4-HB-cysteine) | 3.44 | 2.11 | 1.11 | 1.97 |

### Cluster Demographic Composition

The four clusters showed distinct demographic compositions (Figure 9). Non-Hispanic Black participants constituted approximately 40% of the High combustion cluster, substantially exceeding their representation in the Low exposure (approximately 18%) and Moderate mixed (approximately 15%) clusters. The High combustion cluster was 60.8% male and had the highest mean age (45.3 years). The Children/VOC cluster had the lowest mean age (20.3 years) and was approximately gender-balanced. The low-exposure cluster skewed female (58.1%) and had the highest Non-Hispanic White representation.

**Figure 9:**
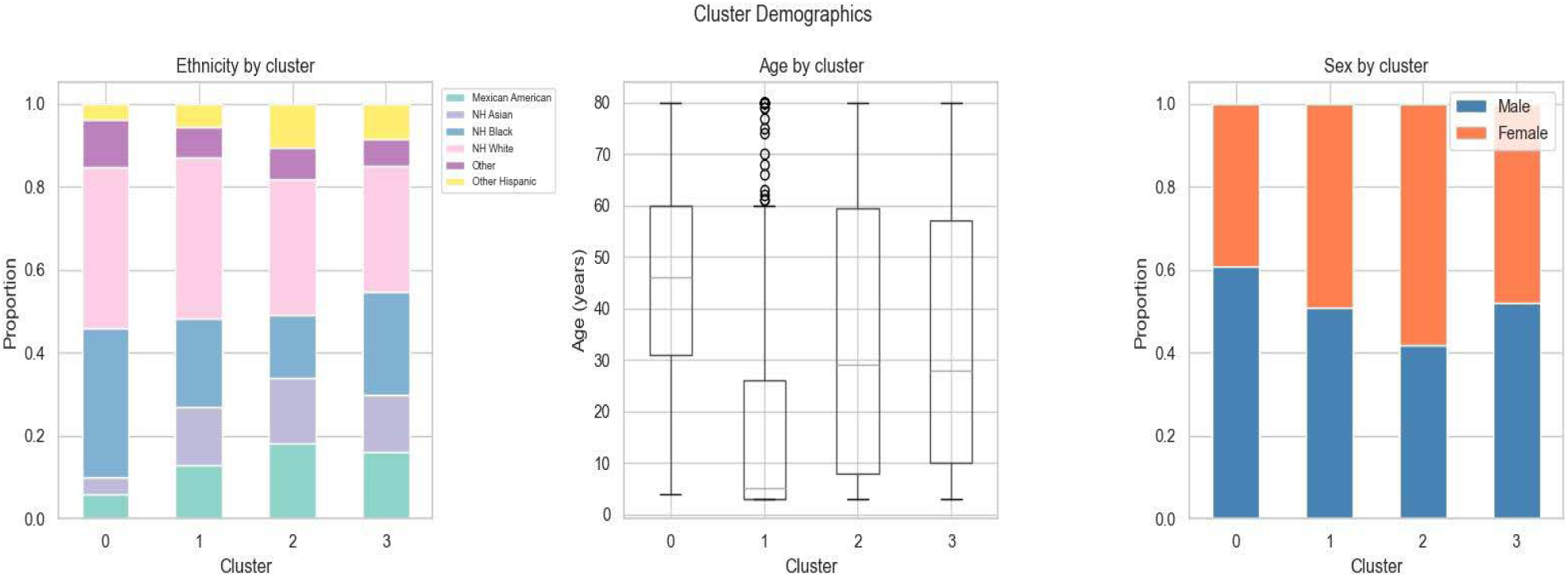
Cluster Demographic Composition. Demographic composition of the four exposure clusters. Left: race/ethnicity proportions by cluster (stacked bars). Centre: age distributions by cluster (box plots). Right: sex proportions by cluster (stacked bars). Non-Hispanic Black participants are disproportionately represented in the High combustion cluster. The Children/VOC cluster has a strikingly younger age profile reflecting paediatric household exposure patterns.

**Table 2:**
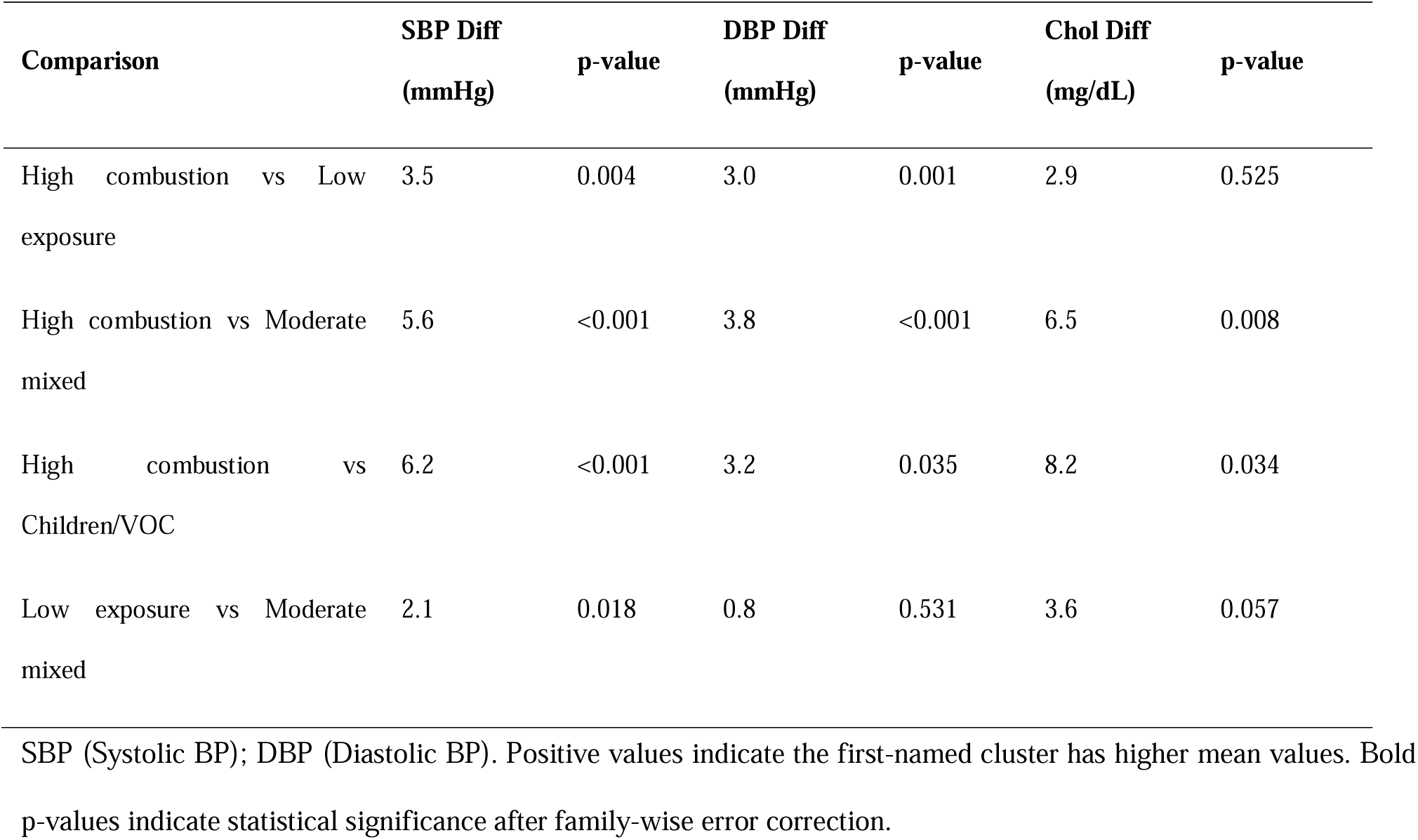
Tukey HSD post-hoc pairwise comparisons between clusters for systolic BP, diastolic BP, and total cholesterol.

### Clinical Outcomes by Cluster

Boxplot examination of the three clinical outcomes confirmed visible between-cluster differences, with the High combustion cluster showing consistently elevated distributions for all three variables and the Children/VOC cluster showing lower values reflecting its younger age profile (Figure 10). One-way ANOVA confirmed statistically significant between-cluster differences for all outcomes: systolic BP (F=11.92, p<0.0001), diastolic BP (F=7.59, p<0.0001), and total cholesterol (F=5.09, p=0.0016).

**Figure 10:**
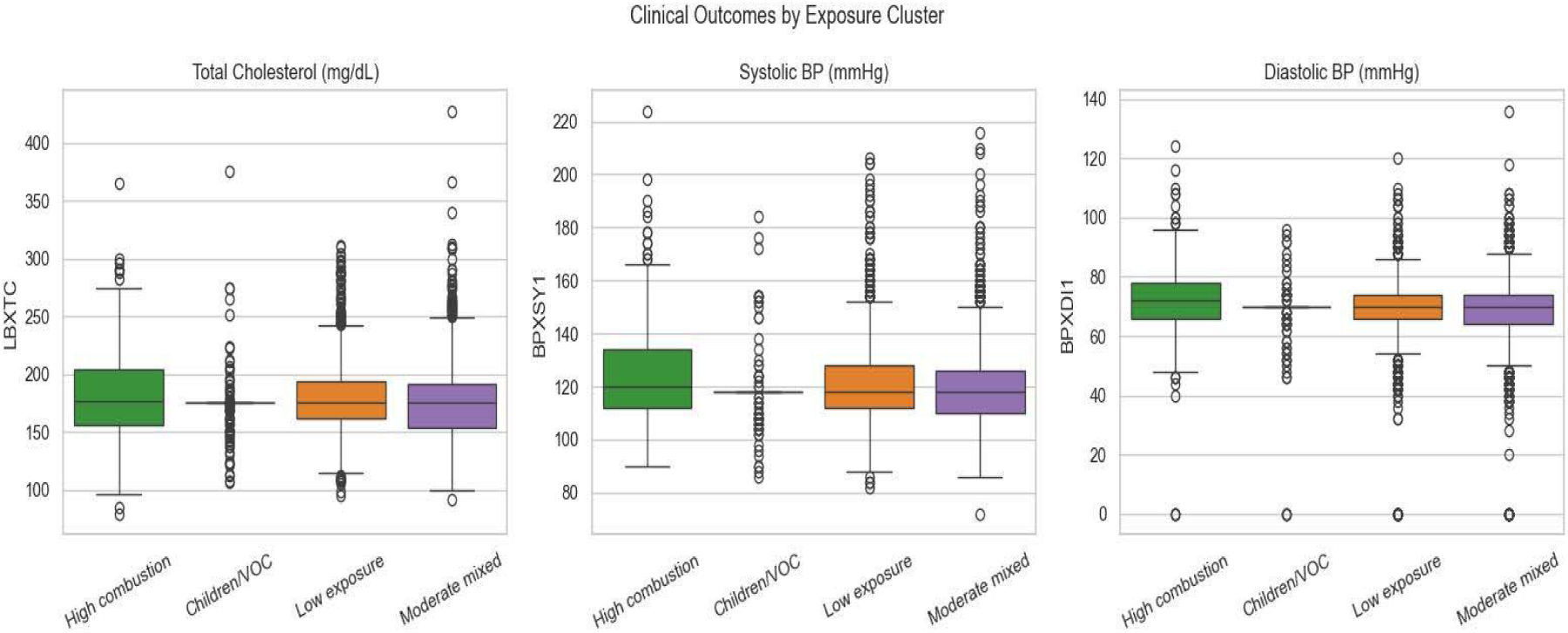
Clinical Outcomes by Exposure Cluster. Box plots of total cholesterol (left), systolic blood pressure (centre), and diastolic blood pressure (right) by exposure cluster. The High combustion cluster (green) shows the highest medians for all three outcomes. The Children/VOC cluster (blue) shows the lowest values, attributable to its younger age composition.

Tukey HSD post-hoc analysis demonstrated that the High combustion cluster had significantly higher systolic BP than both the Low exposure (mean difference 3.5 mmHg, p=0.004) and Moderate mixed clusters (mean difference 5.6 mmHg, p<0.0001). Diastolic BP was similarly elevated in the High combustion cluster relative to Low exposure (3.0 mmHg, p=0.001) and Moderate mixed (3.8 mmHg, p<0.0001). For total cholesterol, High combustion differed significantly from the Moderate mixed group (6.5 mg/dL, p=0.008) and Children/VOC group (8.2 mg/dL, p=0.034).

### Survey-Weighted Prevalence Estimates

Survey-weighted hypertension prevalence in the High combustion cluster was 39.3% (95% CI 37.2-41.4%), representing a 10.6 percentage point excess relative to the Low exposure reference group (28.7%, 95% CI 21.9-35.5%). High combustion cluster hypertension prevalence was 31.2% higher in relative terms than the Moderate mixed cluster (28.9%, 95% CI 22.1-35.7%). Comparison of weighted versus unweighted estimates confirmed robustness: the High combustion hypertension estimate shifted minimally from 40.0% (unweighted) to 39.3% (weighted), while estimates for Low exposure and Moderate mixed clusters widened appropriately with survey weighting (Figure 13).

**Figure 11:**
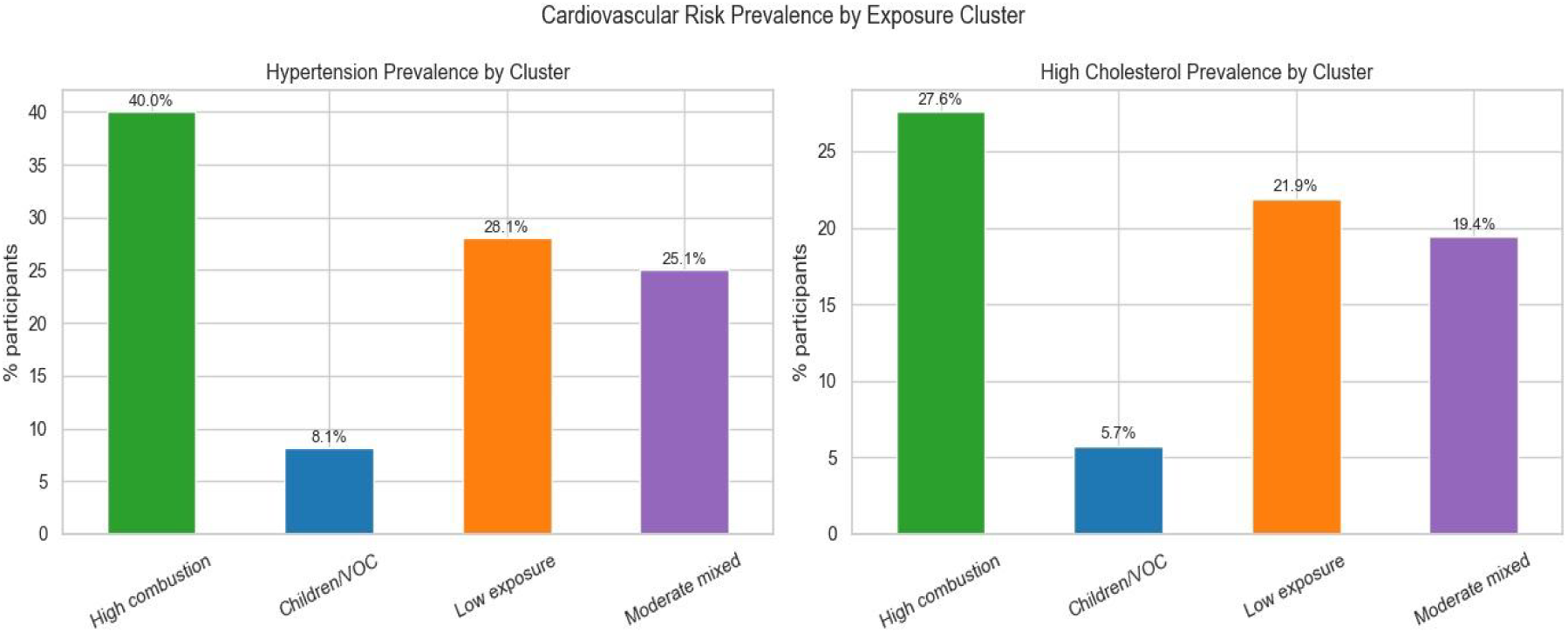
Cardiovascular Risk Prevalence by Cluster. Survey-unweighted prevalence of hypertension (left) and high total cholesterol (right) by exposure cluster. Percentages indicate cluster-level prevalence. The High combustion cluster shows the highest rates for both outcomes.

**Figure 12:**
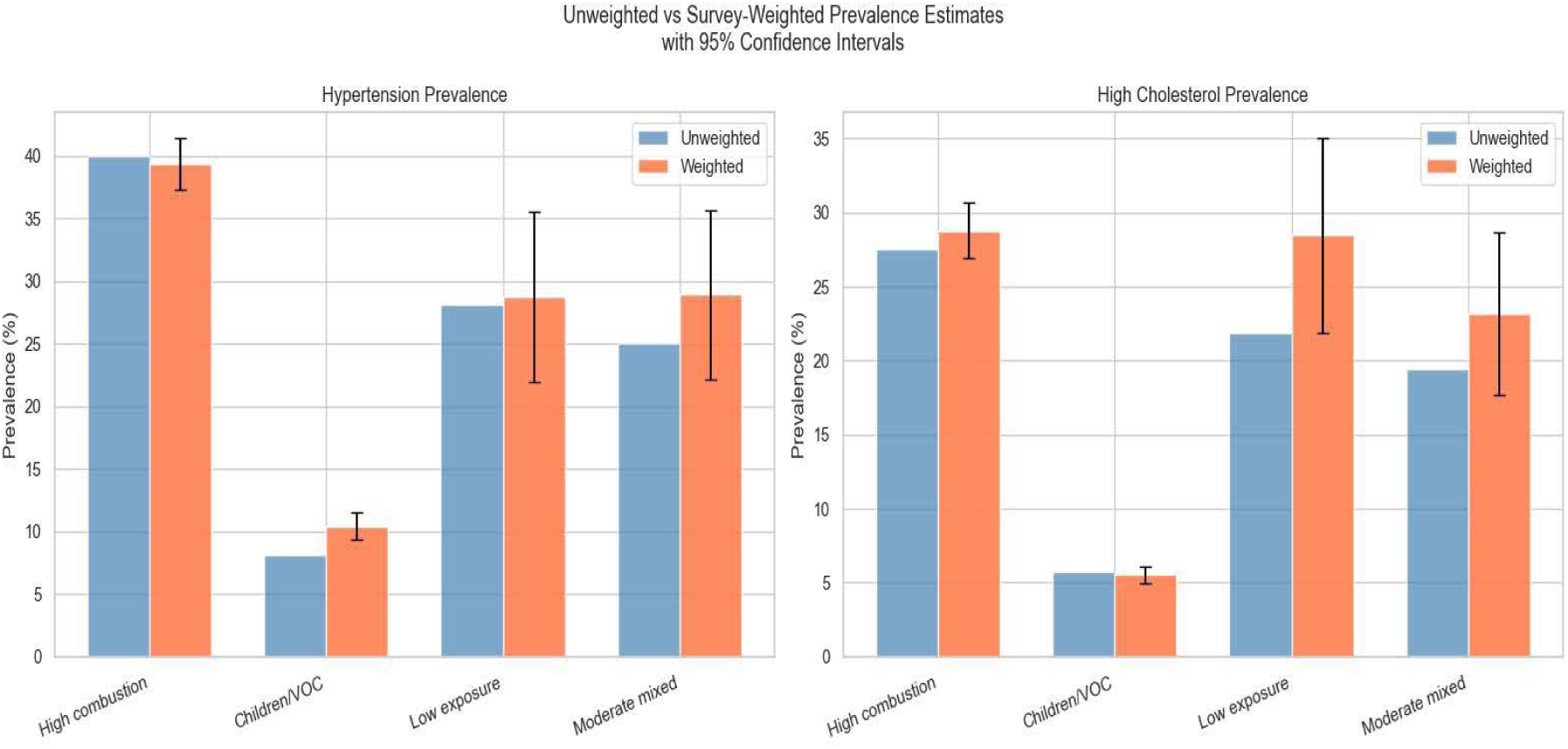
Unweighted vs Survey-Weighted Prevalence Estimates. Comparison of unweighted (blue) and survey-weighted (coral) prevalence estimates with 95% confidence intervals (error bars) for hypertension (left) and high total cholesterol (right) by cluster.

**Figure 13:**
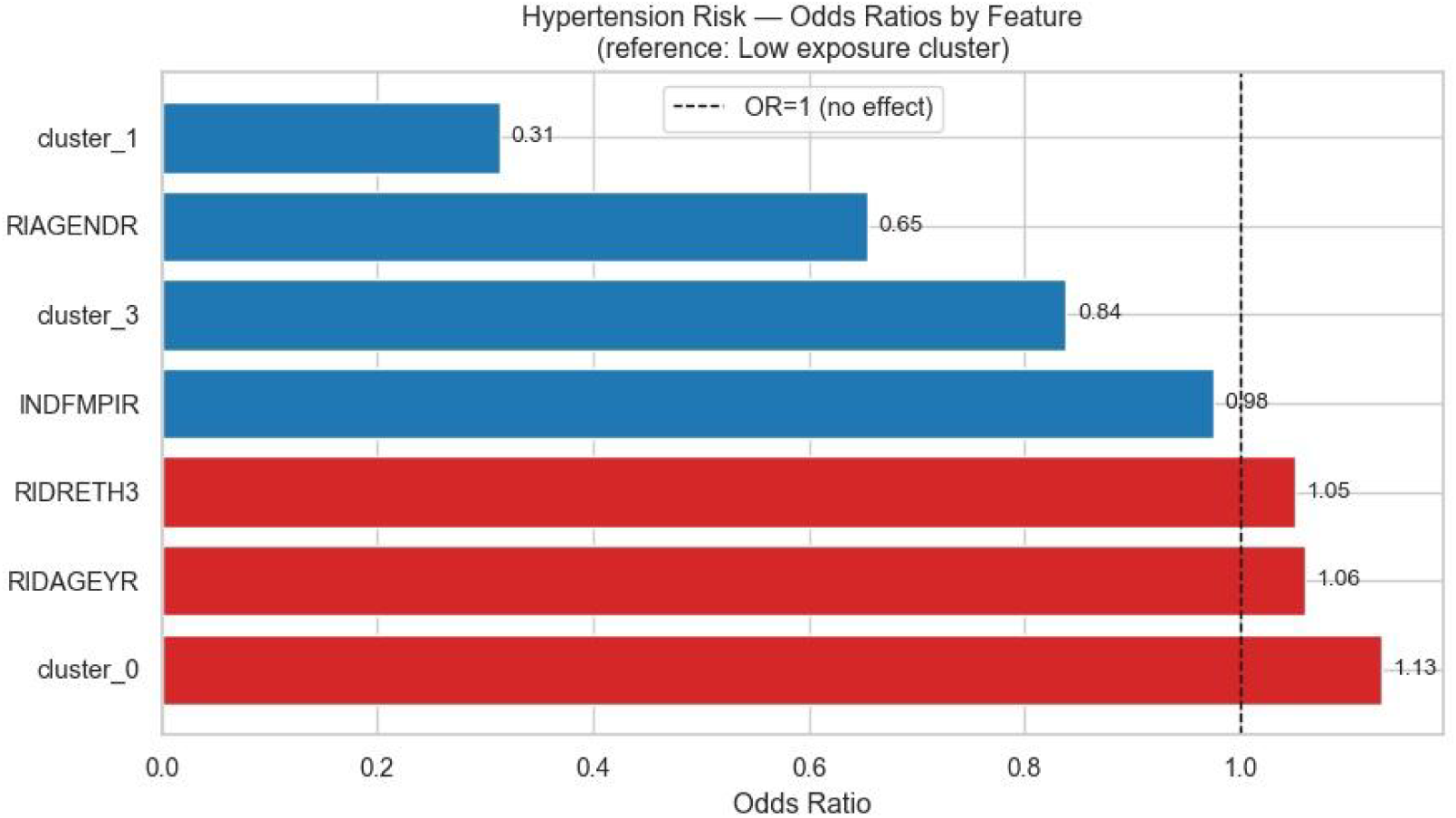
Hypertension Risk: Odds Ratios Forest Plot. Forest plot of weighted multivariable logistic regression odds ratios for hypertension. Dashed vertical line at OR=1.0 indicates the null. Red bars (OR>1) indicate risk factors; blue bars (OR<1) indicate protective associations. The High combustion cluster is the only exposure-related predictor on the risk side of the null, with an OR exceeding those of age and race/ethnicity.

**Table 3:** Survey-weighted cardiovascular risk estimates by exposure cluster.

| Cluster | n | Weighted<br>N | SBP mmHg<br>(95% CI) | DBP mmHg<br>(95% CI) | Total Chol mg/dL<br>(95% CI) | HTN %<br>(95% CI) | High Chol %<br>(95% CI) |
| --- | --- | --- | --- | --- | --- | --- | --- |
| High combustion | 370 | 5,060,825 | 123.0 (122.9-123.2) | 71.8 (67.4-76.1) | 183.3 (175.3-191.3) | 39.3 (37.2-41.4) | 28.8 (26.9-30.6) |
| Children/VOC | 209 | 1,991,973 | 118.5 (117.9-119.1) | 68.8 (66.3-71.3) | 172.3 (169.6-175.1) | 10.4 (9.3-11.5) | 5.5 (5.0-6.1) |
| Low exposure (ref) | 1,019 | 12,765,788 | 120.5 (118.6-122.3) | 69.5 (68.3-70.6) | 183.3 (176.8-189.8) | 28.7 (21.9-35.5) | 28.4 (21.9-35.0) |
| Moderate mixed | 1,381 | 17,007,312 | 120.2 (118.1-122.4) | 69.8 (68.8-70.8) | 178.9 (171.5-186.3) | 28.9 (22.1-35.7) | 23.1 (17.6-28.6) |

HTN: hypertension (SBP ≥ 130 or DBP ≥ 80 mmHg per ACC/AHA 2017). High Chol: total cholesterol ≥ 200 mg/dL. Weighted N estimated using WTMEC2YR/3. 95% CI computed using jackknife variance estimation with SDMVPSU and SDMVSTRA.

### Multivariable Weighted Logistic Regression

After adjustment for age, sex, race/ethnicity, and income, High combustion cluster membership was independently associated with 38.4% higher odds of hypertension relative to the Low exposure reference group (OR 1.38; Table 4; Figure 13). This effect exceeded the per-year age effect (OR 1.06 per year) and the race/ethnicity coefficient (OR 1.06), establishing multi-chemical exposure pattern as the dominant modifiable predictor in the model. The five-fold cross-validated AUC of 0.849 (SD 0.017) indicated strong and stable model discrimination.

**Table 4:** Weighted multivariable logistic regression odds ratios for hypertension.

| Predictor | Weighted<br>OR | Direction | Clinical Interpretation |
| --- | --- | --- | --- |
| High combustion cluster<br>(ref: Low exposure) | 1.38 | Risk | 38.4% higher odds of HTN |
| Age (per year) | 1.06 | Risk | 6.0% higher odds per additional year |
| Race/ethnicity<br>(RIDRETH3) | 1.06 | Risk | Small independent ethnic gradient |
| Moderate mixed cluster | 1.05 | Risk | Slight elevation vs reference |
| Poverty-income ratio | 0.93 | Protective | Higher income marginally protective |
| Children/VOC cluster | 0.56 | Protective | Substantially driven by younger age |
| Female sex | 0.48 | Protective | 52% lower odds vs male (ref) |

Reference group: Low exposure cluster. Model adjusted for age, sex, race/ethnicity (RIDRETH3), and poverty-income
ratio (INDFMPIR). 5-fold cross-validated AUC: 0.849 (SD 0.017). HTN: hypertension.

## DISCUSSION

This study presents, to our knowledge, the first application of Uniform Manifold Approximation and Projection (UMAP) combined with Gaussian Mixture Model clustering to integrated urinary PAH and VOC biomarker data in a nationally representative US population. By leveraging a multi-chemical exposome framework, we identified four distinct exposure clusters that were both chemically interpretable and demographically patterned and demonstrated that membership in a high combustion exposure cluster is associated with higher odds of prevalent hypertension after adjustment for major sociodemographic factors. The pmodel’s redictive performance oAUC 0.849) further indicates that these multi-chemical exposure patterns capture variation in cardiovascular risk not fully explained by conventional covariates.

Our findings broadly align with prior literature linking individual PAH and VOC biomarkers to cardiovascular outcomes, while extending this work by capturing the combined effects of correlated exposures. Previous NHANES-based analyses have reported modest positive associations between individual urinary PAH metabolites and cardiovascular disease endpoints, including hypertension and dyslipidemia□. Similarly, systematic reviews and meta-analyses have demonstrated consistent associations between urinary PAH metabolites and elevated blood pressure□. The stronger association observed in the present study (OR 1.38) likely reflects the multi-chemical nature of the exposure construct. By modeling the full co-exposure structure rather than individual analytes in isolation, our approach more closely approximates the cumulative biological burden experienced in real-world environmental settings, where exposures rarely occur independently.

The biological plausibility of these findings is supported by well-characterized mechanistic pathways linking combustion-derived pollutants to cardiovascular dysfunction. PAHs are known to activate the aryl hydrocarbon receptor, generating oxidative stress and impairing endothelial function through reduced nitric oxide bioavailability and activation of pro-inflammatory signaling pathways such as NF-κB□. Concurrently, VOC metabolites associated with benzene and acrolein exposure contribute to systemic inflammation, hematopoietic disruption, and prothrombotic states□. The co-occurrence of these mechanistically distinct but convergent pathways within the high combustion cluster suggests that cardiovascular risk may arise from the combined effects of multiple co-occurring chemical exposures. This reinforces the limitation of single-pollutant models, which are not designed to capture such interaction effects and may underestimate the true impact of environmental mixtures. An important dimension of this analysis is the observed racial and ethnic patterning of exposure clusters. Non-Hispanic Black participants were disproportionately represented in the high combustion cluster, consistent with a substantial body of literature documenting structural inequities in environmental exposure^12,13^. Prior work has demonstrated that communities of color are systematically exposed to higher levels of combustion-related pollutants due to historical and ongoing patterns of residential segregation, industrial zoning, and the placement of transportation infrastructure¹³. Our findings extend this literature by showing that these disparities are reflected not only in ambient exposure estimates but also in internal biomarker profiles and are associated with measurable differences in cardiovascular risk at the population level. While the present cross-sectional analysis does not permit causal inference, the observed clustering patterns are consistent with the hypothesis that structural environmental inequities contribute to disparities in cardiovascular health.

The identification of a distinct Children/VOC cluster provides additional insight into exposure heterogeneity across the population. This cluster, characterized by younger age and relatively elevated VOC metabolite concentrations with lower PAH levels, is consistent with exposure patterns expected from household or secondhand combustion sources rather than occupational or traffic-related combustion. Although this group exhibited lower absolute cardiovascular risk, this is attributable to age rather than absence of exposure-related effects. Emerging longitudinal evidence suggests that early-life exposure to combustion-related pollutants may influence long-term cardiovascular risk trajectories through developmental and epigenetic mechanisms, which would not be detectable in a cross-sectional framework. As such, this cluster highlights the importance of life-course approaches in exposome research.

Several limitations were considered in interpreting these findings. First, the cross-sectional design precludes causal inference; the observed associations should be interpreted descriptively rather than causally. Second, residual confounding is possible, particularly regarding tobacco use, dietary factors, physical activity, and proximity to pollution sources. Third, urinary biomarkers reflect recent exposure over a short time window and may not fully capture cumulative long-term exposure, potentially leading to exposure misclassification that biases result toward the null. Finally, the analytic sample size, determined by the NHANES subsample design, limits statistical power for finer subgroup analyses. Despite these limitations, this study has several notable strengths. It uses a nationally representative dataset with standardized biomarker measurements, applies a multi-chemical analytical framework aligned with the exposome paradigm, and incorporates appropriate survey weighting and variance estimation to enable population-level inference. The integration of non-linear dimensionality reduction with probabilistic clustering enables the identification of exposure subgroups that would not be detectable with conventional linear methods. The strong and stable predictive performance observed across cross-validation further supports the robustness of the identified exposure patterns.

## CONCLUSIONS

Unsupervised multi-chemical exposome clustering of 25 urinary PAH and VOC biomarkers identified four cardiovascularly distinct subpopulations in a nationally representative US adult population. The High combustion cluster, estimated to represent 5.1 million Americans, exhibited a weighted hypertension prevalence of 39.3% (95% CI: 37.2–41.4%) and was associated with 38.4% higher odds of prevalent hypertension after demographic adjustment. Non-Hispanic Black participants were disproportionately represented in this highest-risk cluster, consistent with structural inequities in combustion-source pollution burden. These findings demonstrate that multi-chemical, data-driven approaches provide greater insight into population cardiovascular risk architecture than single-chemical frameworks and offer a meaningful strategy for characterizing real-world environmental exposures. More broadly, they highlight the importance of incorporating environmental mixtures and structural determinants of exposure into cardiovascular epidemiology and support the public health relevance of reducing combustion-related exposures in disproportionately affected communities.

## DECLARATIONS

### Ethical Standards Disclosure

Not Applicable

### Consent for Publication

Not applicable.

## Data Availability

All data analysed in this study are publicly available from the National Center for Health Statistics (NCHS) at the Centers for Disease Control and Prevention (CDC). The NHANES 2017-2018 datasets used include PAH_J, UVOC_J, SSUVCM_J, P_SSUVCM, DEMO_J, BPX_J, TCHOL_J, and HEPBD series files, all freely accessible at https://wwwn.cdc.gov/nchs/nhanes/continuousnhanes/default.aspx?BeginYear=2017. Analysis code is publicly available at https://github.com/anthoniooladimeji11-coder/nhanes-exposome-cvd-risk

## Acknowledgments

Not applicable.

## Availability of Data and Materials

The dataset(s) supporting the conclusions of this article is(are) available in the GitHub repository, [https://github.com/anthoniooladimeji11-coder/nhanes-exposome-cvd-risk].

## Author’s Contributions

AOG conceptualized the study, designed the study, performed the literature search, and screened and reviewed articles for inclusion. AOG and BIO wrote the first draft of the manuscript. AOG did the data analysis. AOG, BIO, DAO, ATV, and OJA did the final literature review, proofreading, and copyediting. AOG, BIO, DAO, ATV, and OJA reviewed the manuscript and approved it for submission.

